# Navigating the global egg shortage: a comprehensive study of interconnected challenges

**DOI:** 10.1101/2024.03.13.24304250

**Authors:** K Gunjan, Ramendra Pati Pandey, Himanshu, Riya Mukherjee, Chung-Ming Chang

**Affiliations:** Graduate Institute of Biomedical Sciences, Chang Gung University, No. 259, Wenhua 1st Road, Guishan Dist. Taoyuan city 33302, Taiwan (R.O.C); Master & Ph.D. Program in Biotechnology Industry, Chang Gung University, No. 259, Wenhua 1st Road, Guishan Dist. Taoyuan city 33302, Taiwan (R.O.C); Department of Medical Biotechnology and Laboratory Science, Chang Gung University, No. 259, Wenhua 1st Road, Guishan Dist. Taoyuan city 33302, Taiwan (R.O.C); School of Health Sciences and Technology (SoHST) UPES, Bidholi, Dehradun, 248007 Uttarakhand, India

**Keywords:** Egg shortage, Poultry supply chain, Influenza, Economic crisis, One Health surveillance

## Abstract

Poultry eggs are a critical source of protein, vitamins, and minerals for people worldwide; facing the current global egg shortage is a significant concern. The shortage results from various factors, including avian flu outbreaks, changes in consumer demand, and supply chain disruptions caused by the COVID-19 pandemic. The economic crisis caused by the pandemic has also impacted the availability and affordability of eggs, particularly in low-income countries. The global egg shortage has implications for public health, particularly for vulnerable populations who rely on eggs for essential nutrients. One Health, an approach that recognizes the interconnectedness of human, animal, and environmental health, provides a useful framework for understanding and addressing the egg shortage. One Health approach to the egg shortage involves collaboration between agriculture and environmental sectors to address the root causes of the lack and ensure the sustainable production and distribution of eggs. Addressing the global egg shortage requires a multifaceted approach considering the complex social, economic, and environmental factors. One Health perspective offers a way to understand and address the interconnected factors contributing to the shortage to ensure access to affordable, nutritious eggs for all in a healthy way.

## INTRODUCTION

Poultry eggs and meat are the most abundant and primary animal protein sources in the human diet. The egg is regarded as a low-cost, entirely healthy food and a nutritional powerhouse. Both egg whites and yolk are high in protein. It accounts for around 12.6% of an egg’s consumable portion. A, B, E, and K vitamins are also present in eggs. Additionally, eggs include significant amounts of selenium, an antioxidant essential for thyroid, immunological, and mental health as well as zinc, iron, vitamin D, B6 and B12 (Wang et al., 2017). The poultry industry is expanding and becoming more industrialized in many world regions. Growth has been driven by an expanding population, increased purchasing power, and urbanization (View of Major Problems and Challenges of Egg Production and Marketing in Bangladesh, n.d.).

Throughout the previous two decades, the worldwide chicken and egg trade has increased, and poultry meat and egg remain the most traded animal protein(*AGRIS*, n.d.). In the poultry sector, egg production is being impeded by a major global scarcity. The cost of eggs and egg products is growing due to Influenza, lack of supply, an increase in the cost of fuel, feed, and packing, and the manufacturing cost, which are the main factors which lead to running short of eggs in several countries(Egg Shortages and Price Increases Caused in Part by Bird Flu Outbreak - The New York Times, n.d.). The increased cost of eggs results in a price increases for food and confectionery products including eggs as a part of the product. Buyers will face additional challenges as food prices rise. Since October 2021, about 21 million poultry cases have been reported to the World Organization for Animal Health (WOAH, formerly known as the OIE) in various parts of the world(Egg Prices on the Rise: The Effects of Animal Diseases – WOAH – World Organisation for Animal Health, n.d.). Although avian flu is not a new threat, wild birds don’t typically get sick from the virus, but the current circulating strain seems more virulent(Blagodatski et al., 2021). Highly pathogenic avian influenza (HPAI) or “bird flu” is the chief culprit; it can spread swiftly from flock to flock and is fatal in hens. A global pandemic of HPAI viruses of the H5 and H7 subtypes is a worldwide public health problems (Li et al., 2014) and a significant economic loss in the chicken sector(Otte et al., 2008). The avian flu outbreak is the worst in the history of the United States, United Kingdom (UK), and Europe the and also 27 European countries (EU). For the first time, it has across the Atlantic, reaching Canada and the United States (USA), according to the Global food consumer form(Worldwide Egg Shortage Due to Several Reasons Raises Prices in Global Supermarkets - Global Food Consumers Forum, n.d.).

The price and shortage of eggs can have a significant impact on a nation’s economy, particularly in countries where eggs are a staple food and a significant source of protein for the population(Henchion et al., 2017). As egg availability declines and costs rise, households that depend on eggs as a primary protein source may find it difficult to purchase enough to meet their nutritional needs. The shortage may severely impact poultry farmers, forcing them to slaughter birds or reduce production. This could result in lower income for poultry producers and, as a result, more economic hardship for their families. Therefore the ongoing crisis in the global egg supply can also be regarded as “eggflation”.

The general concept of One Health (OH) is widely accepted(Mackenzie & Jeggo, 2019). OH underscores the need for a holistic and interdisciplinary approach to address the interconnected challenges of influenza outbreaks and the global egg shortage. The transmission of Influenza viruses between different species, such as birds, pigs, and humans, highlights the interconnectedness of One Health(Debnath et al., 2021). For example, certain strains of Influenza can originate in animals and then jump to humans, leading to outbreaks or pandemics. OH approach can help to address these issues by identifying and addressing the underlying factors that contribute to egg shortages or price increases, such as disease outbreaks in poultry farms or disruptions in the supply chain.

Despite the poultry sector, especially eggs being an integral part of the economy; regrettably, the price increase and scarcity have reduced consumer egg consumption. This study aims to highlight the scarcity of eggs the world is facing now with the OH perspective and economic crisis, which will enable us to explore the potentialities of this sub-sector.

## MATERIALS AND METHOD

### Research Design

This market research study aimed to comprehensively analyze the Egg Food Market, focusing on scarcity of eggs in the 10 countries of the world, in these countries egg inflation is caused due to various factors including avian influenza, cage-free laws, effects of supply and demand and supply chain difficulties. Hence, we employed a quantitative method to collect, analyze, and interpret the egg food related data and provided the market scenario during the period of six years.

### Data Collection and processing

This descriptive research was conducted from January 2018 to March 2023. The study exclusively relied on secondary data, sourced from a diverse array of outlets, such as industry reports, market databases, scholarly literature, governmental publications, and reputable online resources. These data sources were chosen based on their pertinence, trustworthiness, and completeness in offering insights into the Egg Food Market. The study encompassed the analysis of ten countries where the egg shortage was greatly seen: United States, United Kingdom, European Union (EU-27), Taiwan, Japan, India, Australia, Malaysia, and Jakarta.

### Inclusion criteria

To ensure the relevance and reliability of the secondary data, we employed specific selection criteria: 1) studies focusing on the global egg supply chain, including production, distribution, and consumption patterns 2) articles highlighting the impact of avian flu outbreaks on egg production and the subsequent shortage in the global egg supply data covering the past six years (from 2018 to 2023) to provide a current and comprehensive overview of market trends, 3) articles exploring the impact of factors such as avian flu outbreaks, economic factors, and supply chain disruptions on the availability and pricing of eggs, 4) Data sources with a reputation for accuracy, objectivity, and credibility, 5) Data sets that aligned with the research objectives and questions of this study.

## RESULTS

### Economic loss results from a global egg shortage

The egg industry is adaptable and agile, but a confluence of economic and environmental circumstances in 2022 has made things difficult. Farmers will want to meet demand but confront time constraints and cost pressures. The expense of living may rise as egg prices rise, which can affect the purchasing power of the people and lead to inflation. In addition, if a country is reliant on imports of eggs, then a shortage or price increase can create trade imbalances, as the cost of importing eggs will increase. This can lead to a drain on foreign reserves and a weakening local currency. Furthermore, the egg industry is an essential component of the agricultural sector. The shortage of eggs can affect the livelihoods of farmers and suppliers, mainly if they rely on eggs for their income. The price increase can also shift the demand for other food items and affect the production of other agricultural products. Meanwhile, Eggs are a significant source of protein, and a shortage or price increase can lead to malnutrition, particularly among the low-income sections of the population. This can have a long-term impact on the health of the people and can lead to increased healthcare costs. Overall, a scarcity and hike in the price of eggs can result in a ripple effect on the economy, influencing various industries and causing significant harm to underprivileged communities that depend on eggs as a fundamental source of sustenance. The “eggflation” in many nations is illustrated country wise in the Figure 1.

**Figure 1:**
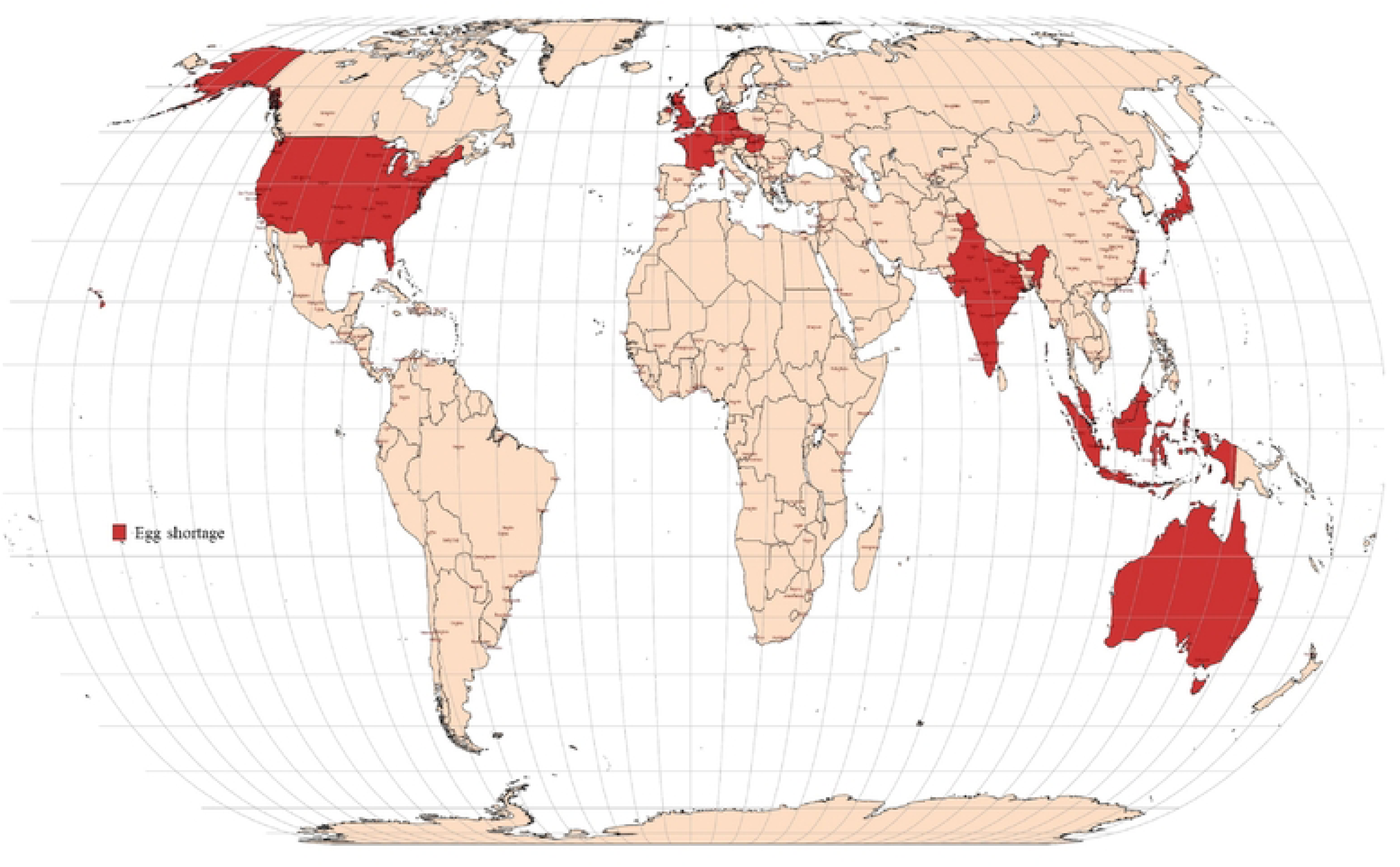
Worldwide egg shortage

### United States

The United States (USA) is facing an exacerbating scarcity of eggs caused by a combination of factors, including the highest avian flu outbreak ever recorded globally and increasing fuel, feed, and packaging costs. According to an article in The “New York Times”, Since the start of 2022, HAI has killed around 58 million commercial and backyard flock birds spanning 47 states, including approximately 44 million egg laying hens in the USA since the epidemic (Egg Shortages and Price Increases Caused in Part by Bird Flu Outbreak - The New York Times, n.d.). Consumer price index, according to the “U.S. Bureau of Labor Statistics”, revised on January 12, 2023, egg prices have increased by up to 59.9% year on year. The average cost for a dozen large Grade A eggs in the USA in January 2022 was $1.929. The average price had risen to $4.823 by January. Nonetheless, it is gradually reducing to $4.221 by February, but it is still high compared to 2022(Bureau of Labor Statistics Data, n.d.).

### United Kingdom

According to data from the British Free Range Egg Producers Association (BFREPA), with about 38 million laying hens across the country, the United Kingdom (UK) is currently 92% self-sufficient in eggs. Flock size had decreased from 44 million to around 36.7 million birds species by November 2022(Egg Producers Continue to Quit on Weekly Basis, Sector Warns - FarmingUK News, n.d.). According to government data, up to September 2022, in the third quarter of 2022, egg packaging plants in the UK packed 213 million dozens of eggs. This represents a 9.6% reduction from the third quarter of 2021 and a 4.1% decline from the second quarter of 2022. The number of eggs processed at packing stations in the third quarter of 2022 declined by 9.6% from the same quarter in 2021(Historical Statistic Notices on UK Egg Production and Prices, 2022 - GOV.UK, n.d.). After seasonal adjustment, the price of eggs in the United Kingdom increased by 8.5% in January 2023 compared to December. While this was a substantial and anticipated drop from December’s commotion-causing 11.1% increase, it did little to mitigate a staggering 79.1% year-over-year growth. (Egg Prices Continue to Climb at Slower Pace alongside Other Food Consumed at Home, CPI Reveals, n.d.).

### European Union (EU-27)

According to an article from Global Food Consumers’ forum, egg prices in the EU have increased by 75% in the previous six months due to supply issues. The 99% of French people who eat eggs this year may eat fewer eggs by up to 10%. “Franceinfo” reports that beginning September 2021, the cost of a box of six eggs has increased by 13 %. Farmers in France have been forced to butcher over 770,000 animals due to the spread of avian flu. As a result, fewer chickens are available to lay eggs, which leads to a decrease in egg production (Worldwide Egg Shortage Due to Several Reasons Raises Prices in Global Supermarkets - Global Food Consumers Forum, n.d.). According to a recent European Commission report released on February 22 2023, grade A egg prices rose to 249.02 EUR/100 kg(*Eggs*, n.d.). According to an article in “The Economic Times”, Egg prices increased the most in the Czech Republic, climbing 85% year on year, followed by two other Central European countries, Hungary (80%) and Slovakia (7%). Germany (79%) and Luxembourg (79%) were on the other end of the spectrum, with both reporting 18% growth. (In the EU’s Inflation Crisis, the Humble Egg Takes the Cake - The Economic Times, n.d.). According to Assosia data, a pack of six Staveley Large Free Range Eggs increased by 18.2% to £1.95 early this year, making a pack 30% more expensive than it was at the start of 2022(Why Are Eggs So Expensive RN?, n.d.).

### Taiwan

Many countries are experiencing an egg shortage due to avian flu outbreaks, and Taiwan is also experiencing a severe egg shortage these days. According to an article in “Taipei Times”, by the end of last year, the number of egg-laying hens in Taiwan had fallen from 45 million to 33 million(Egg Shortage Calls for Cooperation - Taipei Times, n.d.). The NT$5 per kg price increase will raise the wholesale price of duck eggs to a record high of NT$80 per kg. According to a CNA report updated on 5th march 2023, The price of locally sourced eggs in wholesale is set to increase by NT$3, moving from NT$52 (equivalent to US$1.70) to NT$55 per catty (600 grams); it is a new high this month, Simultaneously, the farm gate price will also experience a NT$3 increase, reaching NT$45.5 per catty; initially, it was NT$42.5 starting on Thursday (March 9) (Egg Prices in Taiwan Sizzle to All-Time High | Taiwan News | 2023-03-06 12:53:00, n.d.; Egg Prices in Taiwan to Rise Again as Shortage Continues - Focus Taiwan, n.d.; Expatriate Division, Taipei Economic and Trade Office, Jakarta, Indonesia – News, n.d.). Price increases were implemented for the second time in a month due to an egg supply constraint. According to the most recent Council of Agriculture (COA) statistics, daily egg output is 112,000 packages (200 eggs per box), falling 500,000 to 800,000 eggs short of need. To address the short-term supply shortfall, the COA launched an immediate operation in March to import five million eggs from Australia.

### Japan

An avian influenza outbreak and skyrocketing feed costs are behind a shortage of chicken eggs in Japan that has led to increasing prices. Egg prices are rising due to a lack of supply. According to a “Japan Times” story, the nation experienced its first incident in October 2022, and the disease has spread to more than half of the nation’s prefectures. Moreover, According to economic research firm Teikoku Databank, the wholesale egg price per kilo more than quadrupled to ¥327 in February (Rs. 197) from the previous year, Because of Tori-infuruenza, also known as bird flu, throughout the rest of the world-Japan Times(Explained: After US and UK, Why Japan Is Now Facing Egg Shortage, n.d.). Based on a survey conducted by the agriculture ministry, covering 470 outlets nationwide in February, the average price of a 10-egg carton was 262 yen ($1.92). This marked an 8% increase compared to the previous month and a 25% rise compared to the average February prices in recent years. (Egg Prices on the Rise as 15 Million Birds Culled amid Avian Flu Scare | The Asahi Shimbun: Breaking News, Japan News and Analysis, n.d.). By February 2023, the price of a 10-egg carton had more than doubled, reaching 326 ¥ (High Egg Prices Are Hitting Japan (and Its Pancakes) - Unseen Japan, n.d.).On March 2, the agriculture ministry claimed that 15.02 million birds had to be exterminated across the country. The vast majority of these, 13.85 million, are laying hens.

### India

In India, the Maharashtra animal husbandry department announced that the state is experiencing an egg shortage. As per the report of PTI news, Maharashtra consumes around 2.25 crore eggs per day and can generate 1 to 1.25 crore eggs per day. Hence, the state is experiencing a 1 million egg shortage daily(State Facing Shortage Of Eggs: Official | Aurangabad News - Times of India, n.d.). According to The Associated Press, The average cost of a dozen eggs across the country was $3.59 (293.57) in November, up from $1.72 (140.65) the previous year(This Indian State Is Facing Shortage of 1 Crore Eggs per Day | Mint, n.d.). To manage the infection from the previous year, over 43 million (4.3 crores) out of 58 million (5.8 crores) birds were culled. The prices have increased by about 25-35 per cent in just 3-4 months. To meet the current demand, eggs are being imported from Karnataka, Telangana, and Tamil Nadu.

### Australia

Australia is facing a national egg scarcity, causing prices to rise and retail supplies to be in limited supply. Numerous supermarkets limit the amount of cartons of eggs purchased by each customer. Increased cost of food, employment charges, prices for electricity, and poor weather (winter) are the primary reasons of shortages and price hikes in Australia. Many farmers hesitate to raise output in an uncertain economic environment characterized by rising rates of debt and costs. According to the recent report of Global product prices, the whole average price is 2.95 USD; in March 2023, it rose to USD 4.52(Australia - Eggs - Price, March 2023 | GlobalProductPrices.Com, n.d.). The shortage of eggs in the market is due to reduced production, and the pandemic avian influenza is the leading cause of the price rise.

### Other Asian countries

According to the department of statistics Malaysia (DOSM)(Department of Statistics Malaysia Official Portal, n.d.), the chicken egg price is RM 6.56 per kg and RM 0.46 each in September 2022. According to CNA, Malaysia experienced an egg shortage of 157 million in November, up from 118 million in October. For 30 grade A eggs, it used to cost less than RM11 (S$3.35) a tray. The price then increased to RM13.50- as per the report of Straits Times.

In Jakarta’s, Indonesia’s traditional markets, the cost of chicken eggs increased by 30% in June, reaching 30,000 rupiahs per kg. A comparable rise has happened in Bogor and Bekasi, both in West Java-according to an article in “The Star” and “The Jakarta Post”(Egg Prices Break All-Time High, Industry Blames Bulk Social Aid Purchases - Economy - The Jakarta Post, n.d.; Topic: Indonesia | The Star, n.d.).

### Outbreaks of Avian influenza in poultry birds

Recent global outbreaks of HPAI have curtailed egg supplies in the face of increased demand, fueling recent price rises. It continues to be a source of concern in the business, with marketers struggling to keep sufficient stocks on hand to meet current demand. This has become a concern in many countries, resulting in trade reductions and/or elimination for key importing and exporting countries. As illustrated in figure 2, the EU, USA, and UK are experiencing one of their greatest bird flu crises this year, with each market culling millions of birds (Avian Influenza – WOAH – World Organisation for Animal Health, n.d.) (Adlhoch, Fusaro, Gonzales, Kuiken, Marangon, Niqueux, Staubach, Terregino, Aznar, et al., 2022; Agüero et al., 2023; Sidik, 2022). Domestic poultry throughout Asia and Europe and isolated African outbreaks will be affected from 2018 to 2021(Adlhoch, Fusaro, Gonzales, Kuiken, Marangon, Niqueux, Staubach, Terregino, Guajardo, et al., 2022; Baek et al., 2021; Peyrot et al., 2022; Poen et al., 2019; Verhagen et al., 2021).

**Figure 2:**
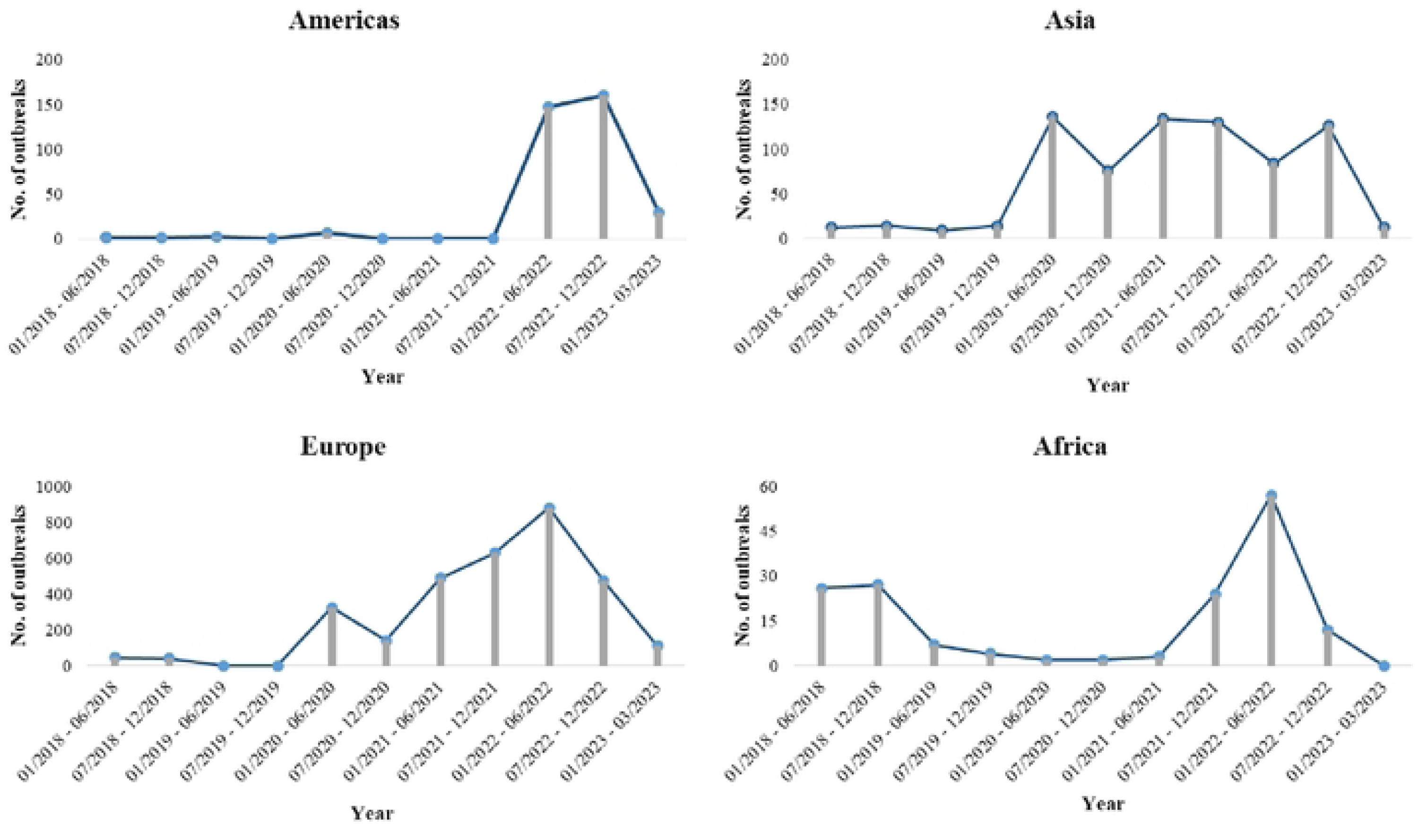
Occurrence of highly pathogenic avian influenza (HPAl) outbreak events in pouluy birds from I January 2018 to 9 March 2023. Accessed fron1 the International Organization for Animal Health documented *(OIE)(Avian Influenza-WOAH* - *World Organisation.for Ani111al Health,* n.d.).

The outbreaks from 2018 to 2021 witnessed the disease and fatality of a substantial number of birds, often surpassing 10,000 individuals. Figure 2 clearly depicts the occurrence of Avian Influenza (AI) in the USA, spanning from 07/2021 to 03/2023, with a noticeable decline in recent times. In Asia, there was a significant surge in outbreaks from 07/2019 to 06/2020, followed by minor fluctuations throughout 2021, culminating in a rapid decrease in 2023. Conversely, Europe experienced a gradual increase in AI outbreaks from 2019 to 2020. In the latter half of 2020, after a slight decrease, there was a substantial uptick in outbreaks from 12/2020 to 06/2022, followed by a marked decline by the end of 03/2023. On the other hand, Africa exhibited a divergent pattern in the figures from 01/2018 to 01/2023. During this interval, there was a sharp increase observed from 01/2022 to 06/2022, succeeded by a pronounced decrease in AI outbreaks until 03/2023.

### One Health

One Health (OH) is a multidisciplinary strategy to preserving human, animal, and environmental health through monitoring, prevention, and mitigation. OH’s major purpose is to attain optimal health and sustainability for humans, animals, and the environment.(Prata et al., 2022; Sharifi & Karamouzian, 2014). Yet, due to the substantial distance between animal and human health, multi-sectoral cooperation in surveillance and controlling new infectious diseases is difficult to achieve. To close this gap, the EU and the USA have contributed money to encourage interdisciplinary research under the OH paradigm, such as research on therapies for new zoonotic diseases and early warning systems of animal risks to humans. Given the importance of the OH strategy in minimizing the public health danger posed by new infectious diseases and current international trends, its implementation through multi-sectoral collaborative initiatives should be discussed regularly. It is crucial to continue investigating the epidemiology of human and avian influenza circulation and hazards by implementing and strengthening an OH strategy for influenza surveillance. In this regard, many organizations worked globally and regionally to combat these epidemics and pandemics.

For influenza, the World Health Organization (WHO), the Food and Agricultural Organization (FAO), and the World Organization for Animal Health are the three international organizations (OIE) primarily responsible for avian flu surveillance and management, and overseeing public health, food safety, and animal health, respectively. (Chien, 2013). The 2010 Tripartite Agreement Concept Note outlined a collaborative strategic direction for the three organizations to pursue and their shared resolve to collaborate more closely in support of countries. FAO, OIE, and WHO reiterated their commitment in 2017, highlighting advances in zoonotic influenza prevention and control while committing to maintain this pace (OIE, FAO and WHO Enlarge Their Collaboration Commitment to Face Health Challenges – WOAH – World Organisation for Animal Health, n.d.). The WHO’s Global Influenza Programme (GIP) establishes global influenza surveillance guidelines. Furthermore, GIP gathers and analyses global virological and epidemiological influenza monitoring data (WHO EMRO | Influenza Surveillance | Influenza | Health Topics, n.d.). Due to the constantly evolving nature of influenza viruses, the Global Influenza Surveillance and Response System (GISRS) is also working to combat this threat. It is a WHO-coordinated network of public health laboratories. GISRS (conducts continuous surveillance of influenza viruses throughout the year. It observes the evolution of these viruses and provides recommendations related to laboratory diagnostics, vaccinations, antiviral susceptibility testing, and risk assessment. Moreover, GISRS serves as a global alert system to identify influenza viruses that could potentially cause pandemics and also has broader implications for emerging respiratory diseases like SARS-CoV and the Middle East respiratory syndrome coronavirus. (MERS-CoV) (Global Influenza Strategy 2019–2030, n.d.) In addition, Global Action Plan for Influenza Vaccines (GAP), it represents an all-encompassing plan to eliminate worldwide shortages. and inequitable access to pandemic influenza vaccinations through three primary approaches: increasing evidence-based seasonal vaccine usage, increasing vaccine production capacity; and promoting research and development of more effective vaccines (Geneva & Switzerland, n.d.). The country-wise surveillance system of OH is also illustrated in Table 1.

## DISCUSSION

The global egg shortage has emerged as a critical concern with far-reaching implications, and it is crucial to understand the factors driving this crisis in egg availability. Our study aims to shed light on the current global egg shortage, examining it through the lenses of the OH perspective and the economic crises unfolding in different countries. Nonetheless, several factors, such as avian influenza outbreaks, legislation promoting cage-free egg production, fluctuations in supply and demand, and complications in the supply chain, have all played a role in the current egg availability crisis.

One significant factor contributing to this shortage is the devastating impact of avian influenza, particularly the highly pathogenic strains (HPAI), on poultry farms worldwide. After introducing severe acute respiratory syndrome coronavirus 2 (SARS-CoV-2), seasonal influenza virus circulation declined dramatically in 2020-21 but increased in 2021-22. This virus has caused the culling of millions of birds, leading to a sharp decline in egg production and subsequently driving up prices due to unrelenting demand (Uyeki et al., 2022).

Furthermore, the shift towards cage-free housing systems has gained momentum, especially in the USA, as it is believed to offer improved living conditions for hens (Lin et al., 2017). By the end of 2019, approximately 80% of table egg-laying chickens were raised in cage systems, commonly known as conventional production. The remaining 20% were produced as specialty eggs, with cage-free eggs being the most prevalent category among them. (Egg Industry Center, 2019). The significant advancement in cage-free production is due to (1) recent regulation compelling farmers to transition from traditional to cage-free production and (2) pledges by huge egg buyers such as McDonald’s, Walmart, Starbucks, and more than 200 restaurants and supermarkets to utilize cage-free products alone by 2025. (Markets Insider 2017). If the commitments made by the participants are kept, around 75% of the laying eggs flock must be cage-free by 2025. Most consumers are only willing to pay an extra $0.30 per dozen for cage-free eggs. Still, the average premium is $1.16 per dozen, demonstrating that only a small fraction of buyers are prepared to pay big profits for the cage-free label, and the market share for cage-free eggs can expand above its current level even at prices as high as $1.00 per dozen. (Lusk, 2019). This transition, driven in part by animal welfare legislation and commitments from major egg buyers, has disrupted the traditional supply chain, with implications for egg prices. The challenge lies in consumers’ willingness to pay a premium for cage-free eggs, which has a ripple effect on the price of conventional eggs.

The delicate balance between supply and demand is another critical element in the egg availability equation. As the world’s population grows, so does the demand for eggs and other protein sources, resulting in the need for increased production. However, the gap between supply and demand has widened, leading to shortages in some regions. To meet their needs, several countries are now importing eggs from others, creating a complex global egg trade network. In India, for example, there is an egg scarcity in Maharashtra; thus, the other states it is importing eggs from Karnataka, Telangana, and Tamil Nadu. Similarly, on March 10, 2023, Taiwan began importing eggs from eight countries, including the USA, Australia, Japan, Brazil, Turkey, Thailand, the Philippines, and Malaysia. Furthermore, according to the Observatory of economic complexity (OCE), the UK is also importing eggs from The USA (£506k or 27.9%), Spain (£84.4k or 48.1%), and Poland (£33.3k or 20.1%) (in November 2022)(Eggs in United Kingdom | OEC - The Observatory of Economic Complexity, n.d.), and Japan is also importing eggs from Hong Kong (484 million), Singapore (14.5 million), Taiwan (4.75 million), and the USA (251,000), and were primarily imported from China (19 million), Taiwan (13.8 million), Germany (4.32 million), and the USA (1.58 million)(Eggs in Japan | OEC - The Observatory of Economic Complexity, n.d.). In addition, in 2020, many countries experienced egg shortages due to the coronavirus disease 2019 (COVID-19) pandemic, which disrupted supply chains and led to changes in consumer demand. At the beginning of the pandemic, panic buying led to a surge in egg demand, causing shortages in some areas. However, as lockdowns and economic disruptions continued, demand for eggs declined, leading to oversupply in some regions and reduced production in others.

Supply chain disruptions have further exacerbated the egg crisis. Complex supply chain structures and shortages of essential supplies and transportation have made it challenging for national egg producers to meet the growing demand, leading to price hikes and supply shortages. Chicken mortality rates have added to this supply/demand conflict, leaving consumers facing higher prices and, at times, empty shelves.

In essence, the production of eggs involves a highly intricate supply chain that extends beyond just birds, feed, water, and land. The pandemic, with its impact on labor, construction costs, and the severe outbreak of HPAI, has exposed unresolved supply chain issues in the egg sector. Rising costs and the recent surge in avian influenza have further aggravated the egg shortage, leaving us with a complex and multifaceted problem that necessitates comprehensive analysis and innovative solutions.

## CONCLUSIONS

The shortage and price increase of eggs can have a cascading effect on the economy, affecting multiple sectors. It can be particularly devastating for low-income sections of the population who rely on eggs as a primary source of nutrition. To address egg shortages, governments may take actions such as importing eggs from other countries, providing subsidies to farmers, or implementing policies to encourage increased production. Additionally, consumers may reduce their egg consumption or switch to alternative protein sources during a shortage. The egg scarcity has demonstrated to egg growers and food sector manufacturers that there is still much opportunity for development and an overall requirement for domesticated and localized supply chain fulfilment techniques. Importing eggs is not a solution for this type of shortage; every country should employ some measures to control influenza as it directly linked with poultry, egg and humans.

## Data Availability

NA

## AUTHOR CONTRIBUTIONS

All authors contributed to writing-original draft preparation, review and editing.

## FUNDING

This research was funded by VtR Inc-CGU (SCRPD1L0221); DOXABIO-CGU (SCRPD1K0131), and CGU grant (UZRPD1L0011, UZRPD1M0081).

## INSTITUTIONAL REVIEW BOARD STATEMENT

Not applicable

## INFORMED CONSENT STATEMENT

Not applicable

## CONFLICTS OF INTEREST

The authors confirm that there are no conflicts of interest.

## Graphical Abstract

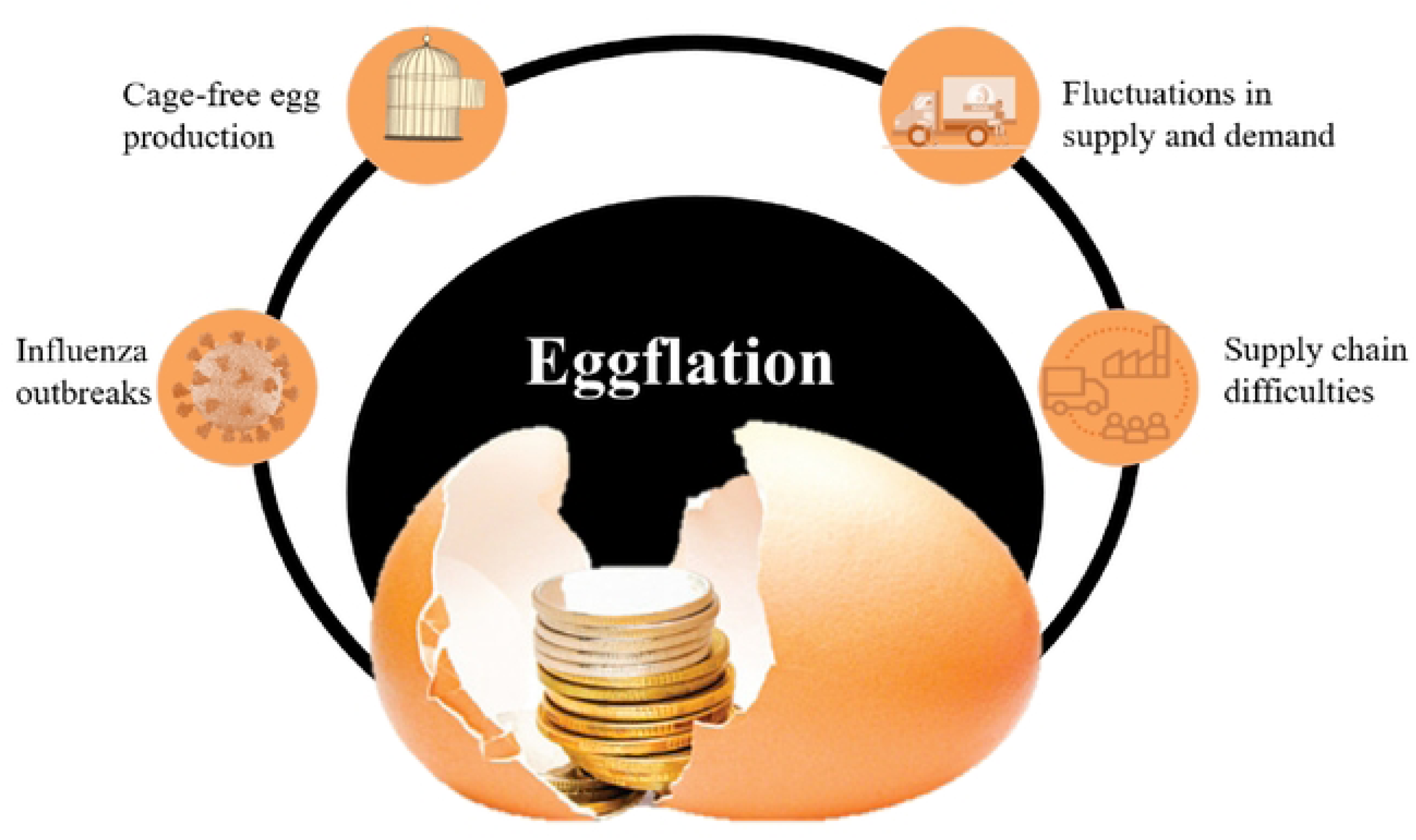

## Notes

### Competing Interest Statement

The authors have declared no competing interest.

### Clinical Trial

NA

### Funding Statement

The author(s) received no specific funding for this work.

